# A Metabolic Epidemic? Prevalence and Sex-Based Disparities of Metabolic Alterations in the Peruvian Population Using Multiple Diagnostic Criteria

**DOI:** 10.1101/2024.08.12.24311881

**Authors:** Víctor Juan Vera-Ponce, Luisa Erika Milagros Vásquez-Romero, Fiorella E. Zuzunaga-Montoya, Joan A. Loayza-Castro, Jhonatan Roberto Astucuri Hidalgo, Carmen Inés Gutierrez De Carrillo

## Abstract

**Introduction:** Metabolic alterations constitute a growing challenge for global public health, with significant implications for cardiovascular morbidity and mortality. Early identification of these alterations, even from the presence of a single component, is crucial for effectively preventing and managing chronic diseases.

**Objective:** To determine the prevalence of metabolic states based on one or more alterations in the Peruvian population and to evaluate possible sex disparities.

**Methods:** An analytical cross-sectional study used data from two Peruvian national databases: Surveillance of Nutritional Indicators by Life Stages (VIANEV) and PERU MIGRANT. Graphs were generated to illustrate metabolic states according to different combinations of diagnostic criteria and even definitions. Bar charts were created to visualize the individual prevalences of each state. Ordinal logistic regression was employed to examine sex disparities and the outcome.

**Results:** The prevalence of metabolic alterations (at least one alteration) ranged from 87.04% to 87.55%, depending on the criteria used. Significant discrepancies were found in the prevalences of hyperglycemia and abdominal obesity according to the different diagnostic criteria applied. In both studies, men showed a lower probability of presenting metabolic alterations than women, regardless of the diagnostic method used.

**Conclusions:** This study reveals a high prevalence of metabolic alterations in the Peruvian population, with notable variations depending on the diagnostic criteria employed. The observed discrepancies underscore the need to re-evaluate and possibly adapt these criteria for the Peruvian and Latin American populations. The identified disparities between sexes suggest the importance of developing differentiated prevention and management strategies.

## Introduction

Metabolic alterations represent a growing challenge for global public health, with significant implications for cardiovascular morbidity and mortality. Traditionally, the focus has been on metabolic syndrome, defined by three or more alterations, or the unhealthy metabolic state, characterized by two or more alterations ^(1)^. However, growing evidence suggests that even a single metabolic alteration can have significant long-term health impacts ^(2)^.

In this context, the present study aims to determine the prevalence of metabolic states based on one or more alterations in the Peruvian population. This more inclusive approach allows for a more comprehensive assessment of metabolic risk in the population, capturing cases of established metabolic syndrome and individuals who might be in the early stages of metabolic alterations ^(3,4)^.

The study utilizes two important databases to achieve this objective: the PERU MIGRANT study and the VIANEV study. These two data sources allow for a more robust and representative assessment of the Peruvian population, encompassing different geographic and sociodemographic contexts. This approach is crucial, given that recent studies have shown that metabolic risk factors can vary significantly among different subpopulations within the same country ^(5)^.

Furthermore, this study distinguishes by sex, exploring disparities in the prevalence of metabolic alterations between men and women. This differentiation is particularly relevant given that recent research has shown significant differences in the manifestation and impact of metabolic alterations according to sex ^(6)^. Understanding these disparities is fundamental for developing more effective and personalized prevention and management strategies.

Finally, by examining the prevalence of metabolic alterations from a broader perspective, this study seeks to provide crucial evidence to inform public health policies and clinical guidelines in Peru. In a context where non-communicable diseases are on the rise, particularly in middle-income countries like Peru, understanding the complete landscape of metabolic alterations is essential for designing effective interventions and allocating resources efficiently.

## Methods

### Study Design

An analytical cross-sectional study used data from two Peruvian national databases: Surveillance of Nutritional Indicators by Life Stages (VIANEV) and PERU MIGRANT. These databases provide valuable information on different aspects of metabolic health in diverse Peruvian populations.

The VIANEV study (2017) ^(7)^ aimed to evaluate the Peruvian population’s nutritional status and associated risk factors at different life stages. The PERU MIGRANT study (2008) ^(8)^ focused on examining the impact of rural-urban migration on cardiovascular risk factors, comparing rural, migrant, and urban populations.

### Populations and Sample

Initially, VIANEV had 1,211 adults between 18 and 59 years old, while PERU MIGRANT worked with 989 participants ≥30. The VIANEV study population was selected through multi-stage probabilistic sampling at the national level. PERU MIGRANT used random probabilistic sampling, focusing on three specific groups (rural, migrant, and urban).

Only participants with complete data for all anthropometric metrics and biochemical analyses considered in the study were included in the present analysis. This restriction in sample selection may have reduced the sample size compared to the original populations of each study, which was considered in the interpretation of results and discussed as a possible limitation of the study.

### Variables

The following variables were evaluated using globally standardized cut-off points:

1. Abdominal obesity (AO) according to Adult Treatment Panel III criteria (AO-ATP) ^(9)^: waist circumference (WC) ≥ 102 cm in men or ≥ 88 cm in women
2. AO according to International Diabetes Federation criteria (AO-IDF) ^(10)^: WC ≥ 94 cm in men or ≥ 80 cm in women
3. Hyperglycemia according to the American Diabetes Association (Hyperglycemia-ADA) ^(11)^: fasting glucose ≥ 100 mg/dL
4. Hyperglycemia according to the World Health Organization (Hyperglycemia-WHO)^(12)^: fasting glucose ≥ 110 mg/dL
5. Obesity/overweight (13): if body mass index (BMI) was ≥ 25 kg/m²
6. Hypertriglyceridemia ^(9)^: if triglycerides ≥ 150 mg/dL
7. Hypercholesterolemia ^(9)^: if total cholesterol ≥ 200 mg/dL
8. Low HDL ^(9)^: if HDL < 40 mg/dL in men and < 50 mg/dL in women
9. Elevated blood pressure ^(9):^ if systolic blood pressure (SBP) ≥ 130 mmHg or diastolic blood pressure (DBP) ≥ 85 mmHg (14)

Four distinct versions of the Metabolic State variable were created from these criteria, each based on a different combination of criteria for AO and hyperglycemia. These variables were coded on an ordinal scale from 0 to 7, where 0 indicates the absence of any alteration of the metabolic state components and 7 indicates the presence of all components. The four combinations were:

- AO-ATP + Hyperglycemia-ADA
- AO-IDF + Hyperglycemia-ADA
- AO-ATP + Hyperglycemia-WHO
- AO-IDF + Hyperglycemia-WHO

The primary independent variable was sex, which was classified as male and female. Additionally, the following covariables were analyzed: age, educational level (none/primary, secondary, and higher), area of residence (urban versus rural), socioeconomic status (non-poor versus poor), physical activity (low, medium, and high), smoking status (current versus non-current), alcohol consumption (evaluated in two ways, according to PERU MIGRANT: heavy drinker versus non-heavy; and VIANEV: excessive versus non-excessive).

### Procedures

For anthropometric measurements, in the VIANEV study, weight was measured with a digital scale (capacity 200 kg, precision 100 g) and height with a wooden stadiometer (precision 1 mm). In the PERU MIGRANT study, weight was recorded with a SECA 940 electronic scale (precision of 0.05 kg) and height with a stadiometer, with an accuracy of 0.1 cm. For WC, VIANEV used a retractable abdominal measuring tape (precision 0.1 cm), while PERU MIGRANT was measured in triplicate at the midpoint between the lower rib and the iliac crest.

For blood pressure measurement, VIANEV used an Omron automatic digital sphygmomanometer, taking two measurements and averaging them; in this case, the dominant arm was identified for blood pressure measurement, and if there was a difference of 20 mmHg in systolic pressure or ten mmHg in diastolic pressure between the first two measurements, a third measurement was taken, recording those that did not present such differences. Additionally, the timing of measurements was specified. PERU MIGRANT measured blood pressure using appropriate cuffs for arm circumference in a seated position, with the right arm supported at chest level. Three measurements were taken with a minimum interval of 5 minutes between them, using an Omron oscillometric device previously validated for the adult population. The average of the last two SBP and DBP measurements was taken for analysis.

Both VIANEV and PERU MIGRANT performed laboratory analyses on venous samples taken in the morning after a minimum 8-hour fast by trained personnel. Fasting glucose was measured in plasma, serum, and whole blood. Additionally, VIANEV used previously calibrated portable glucometers and specified that the automated enzymatic-colorimetric method of coupled reactions at the endpoint was used to determine cholesterol and triglycerides. HDL and LDL determination was performed using the automated direct enzymatic colorimetric method.

### Statistical Analysis

Statistical analysis was performed using the R program for descriptive and inferential analyses, while graphical representations were generated with Python.

A descriptive table of the study population characteristics was elaborated for the descriptive analysis. Age was presented as mean and standard deviation, while other categorical variables were presented as frequencies and percentages. Subsequently, four graphs were generated to illustrate metabolic states according to different combinations of criteria. Additionally, bar graphs were created to visualize the prevalences of each metabolic syndrome component separately.

To examine sex disparities in metabolic state, a bivariate analysis of each type of metabolic state was first presented. Additionally, ordinal logistic regression was used. This model was chosen due to the ordinal nature of the dependent variable (metabolic state), which was categorized from 0 to 7 according to the number of components present. No additional adjustments were made to the model due to the structure of the study variables. It was determined that most potential adjustment variables acted as mediators or colliders in the relationship between sex and metabolic state rather than actual confounding variables. This decision was based on the consideration that sex, as a biological variable, is not a result of other variables in the model but instead influences them.

### Ethical Considerations

Both the PERU MIGRANT and VIANEV databases are freely accessible without restrictions. Therefore, the researchers did not consider it necessary to go through a research ethics committee. However, it is essential to emphasize that both databases were coded to avoid exposing names or any subject identification. Additionally, international standards of research ethics were complied with throughout the research process.

## Results

### Demographic Characteristics – PERU MIGRANT/VIANEV

After applying the selection criteria, PERU MIGRANT worked with 986 subjects and VIANEV with 885 subjects. In both studies, the female sex was predominant (52.84% versus 55.59%), and the number of current smokers was similar (11.16% versus 13.22%). However, regarding other characteristics, there were discrepancies in the wealth index (higher number of poor in PERU MIGRANT) and educational level (higher number with secondary/higher education in VIANEV). They also varied with alcohol consumption (heavy drinker: 9.33% versus excessive consumption: 2.37%) and level of physical activity.

### Consistent Patterns of Metabolic Alterations in Peruvian Populations – PERU MIGRANT/VIANEV

Analysis and comparison of results from the PERU MIGRANT and VIANEV studies, as shown in Figures 1 and 2, reveal notably similar patterns in the distribution of metabolic alterations, regardless of the diagnostic criteria used. Both studies evidence a significant prevalence of multiple metabolic alterations. The prevalence of metabolic alterations (with at least one alteration) ranged from 87.04% to 87.55%, depending on the criteria used.

**Figure 1.**
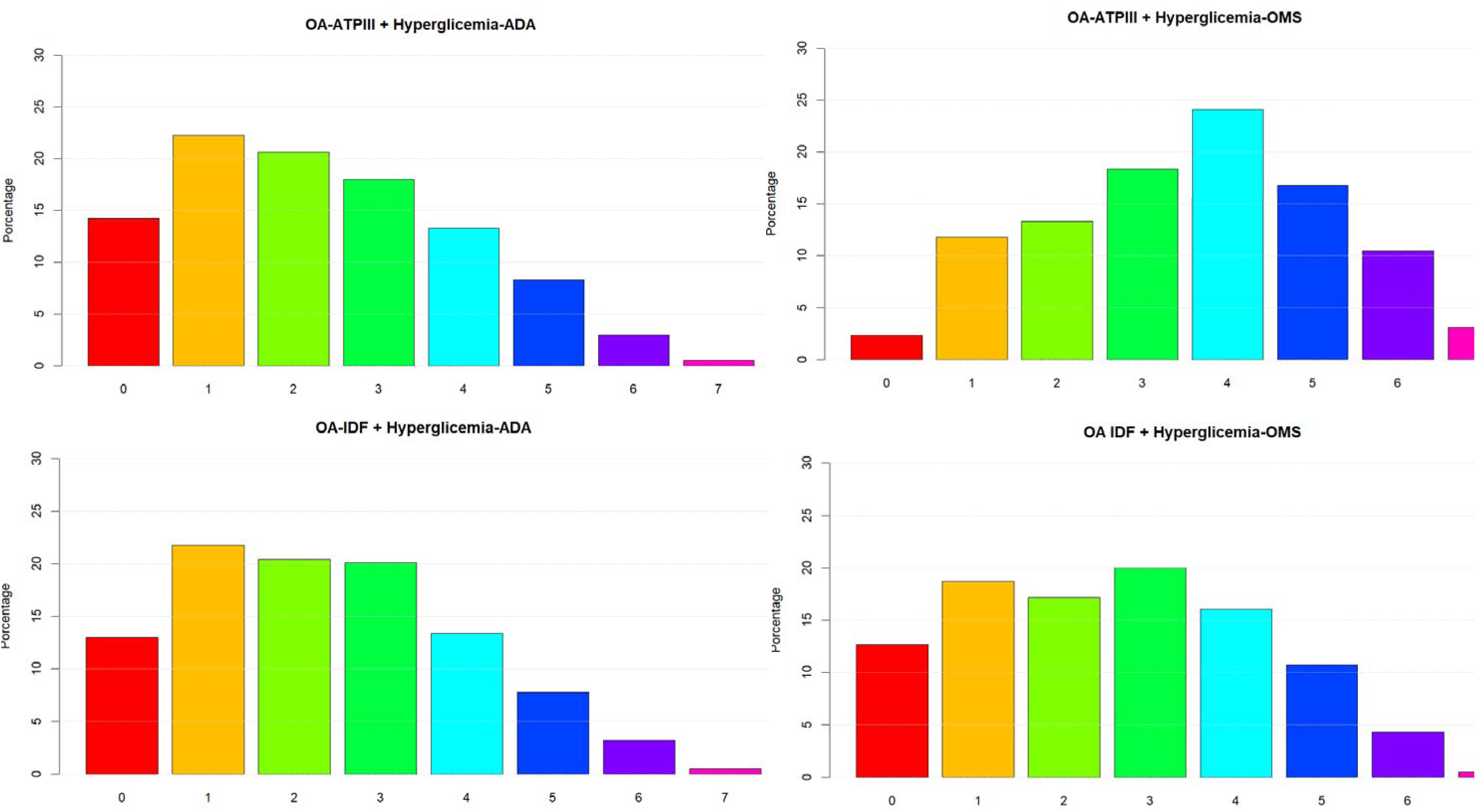
Distribution of metabolic alterations according to different diagnostic criteria in the population of the PERU MIGRANT study.

**Figure 2.**
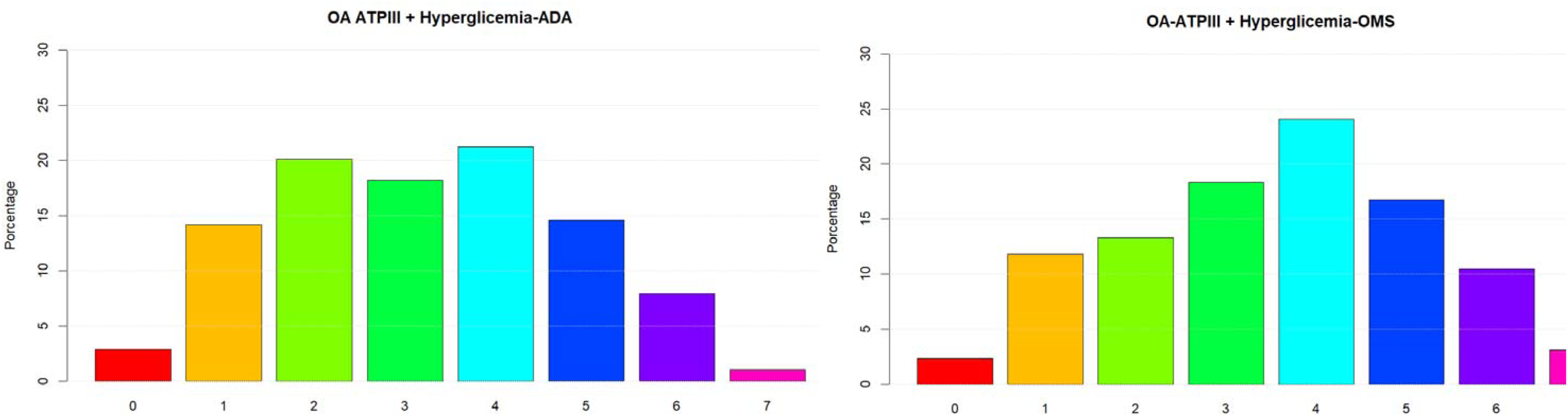
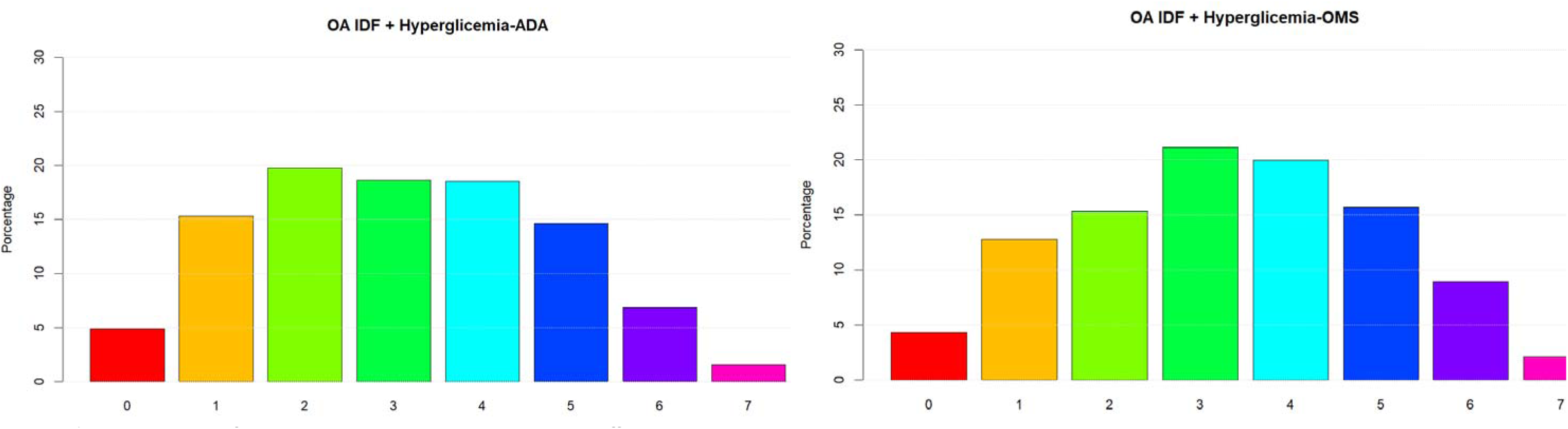
Distribution of metabolic alterations according to different diagnostic criteria in the VIANEV study population.

Notably, the AO-ATP criteria with the OMS hyperglycemia definition consistently result in a higher prevalence of multiple alterations, with a peak at four alterations in both studies. This finding underscores the importance of diagnostic criteria selection in assessing population metabolic health.

Another striking aspect is the low proportion of individuals without metabolic alterations across all criteria combinations in PERU MIGRANT and VIANEV. This raises questions about the adequacy of current cut-off points for the Peruvian population or about the country’s general state of metabolic health.

Both studies exhibit a similar severity gradient, characterized by a gradual decrease in the proportion of individuals as the number of alterations increases. This consistent pattern reinforces the validity of the findings and suggests a distribution of metabolic risk that could be characteristic of the Peruvian population.

### Prevalence of Metabolic Disorders in the Study Populations

Analysis of PERU MIGRANT and VIANEV data revealed consistent patterns in the prevalence of metabolic disorders despite differences in the studied populations. Both studies showed a high prevalence of metabolic alterations, varying according to the diagnostic criteria. You can visualize this in detail in figures 3 and 4.

**Figure 3.**
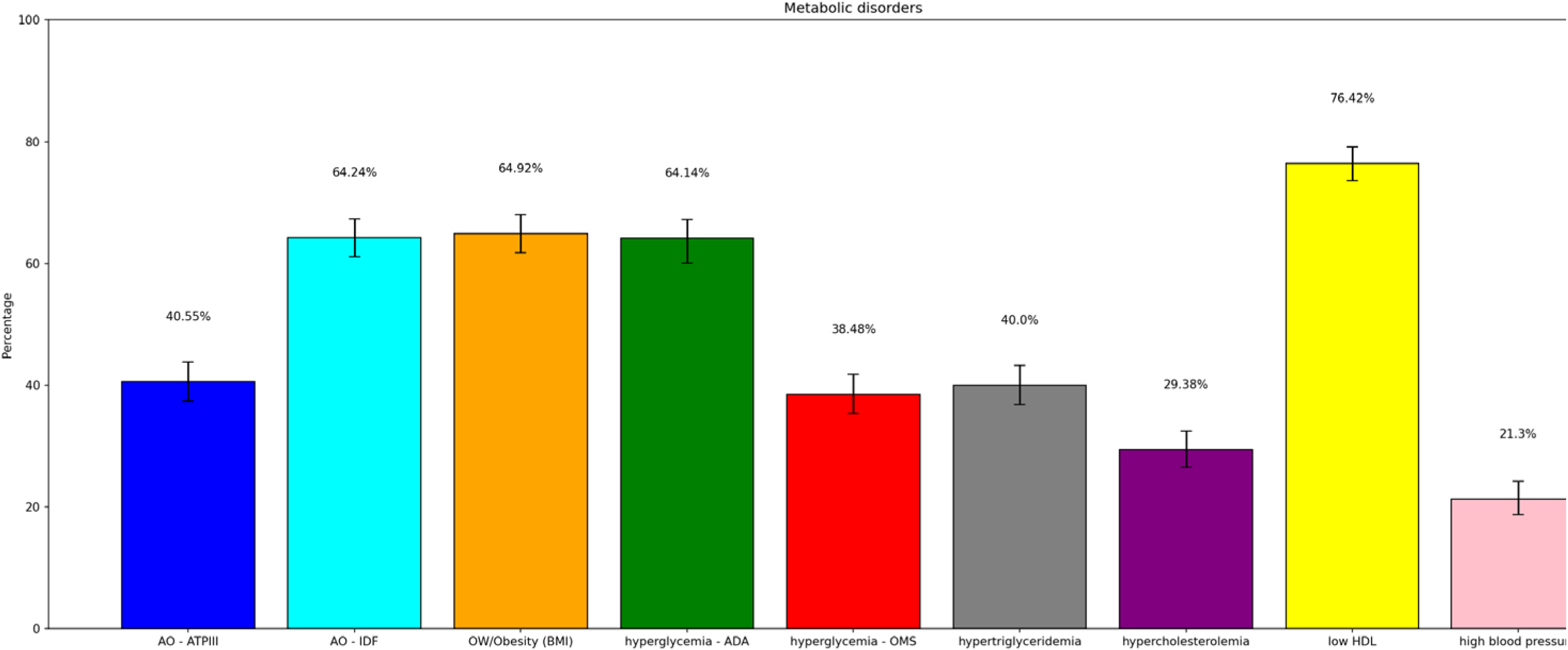
Prevalences of each component of the metabolic state – VIANEV.

**Figure 4.**
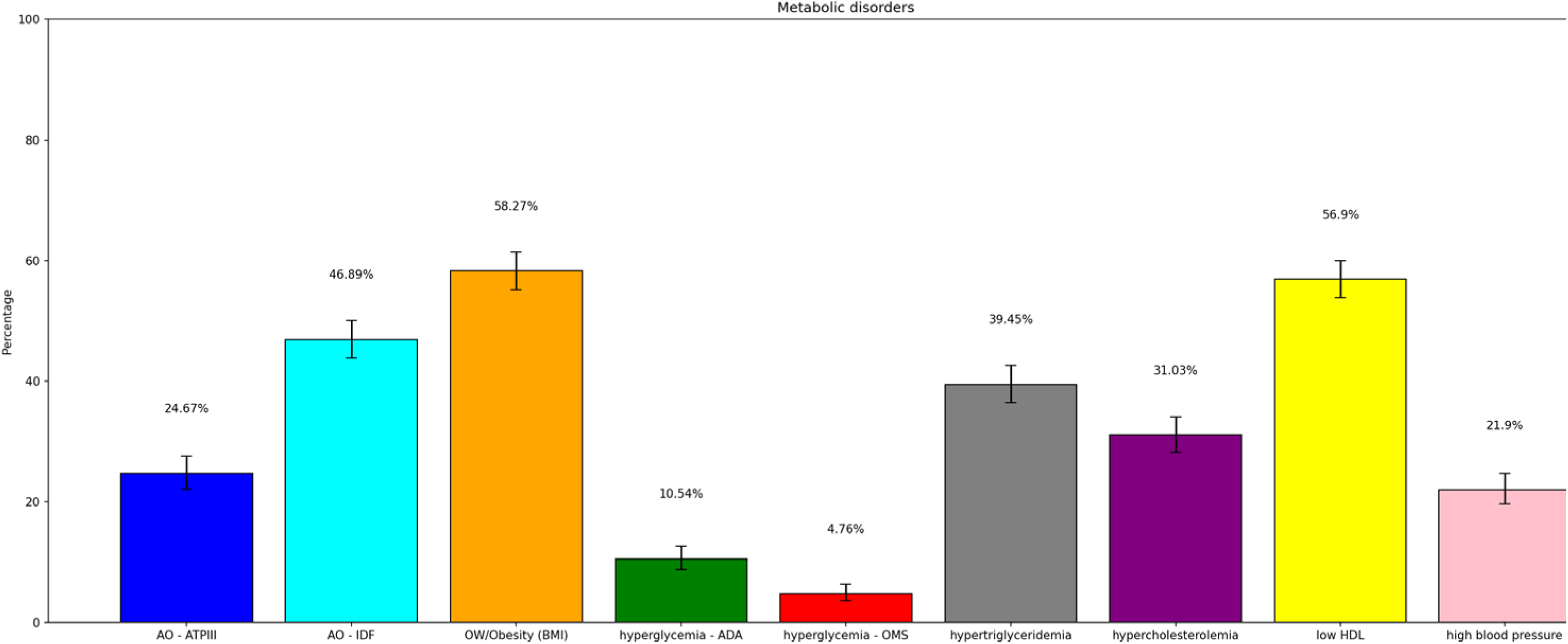
Prevalence of each component of the metabolic state – PERU MIGRANT.

Abdominal obesity presented a high prevalence, with significant differences according to the criteria employed. The IDF criteria resulted in consistently higher prevalences than the ATPIII (64.24% vs 40.55% in PERU MIGRANT, 46.89% vs 24.97% in VIANEV). The prevalence of obesity/overweight by BMI was also high in both studies (64.92% in PERU MIGRANT, 58.27% in VIANEV).

Hyperglycemia showed substantial variation depending on the criterion used. The ADA criteria resulted in notably higher prevalences than the WHO in both studies (64.14% vs 38.44% in PERU MIGRANT, 10.54% vs 4.76% in VIANEV). Among dyslipidemias, low HDL was the most prevalent disorder (76.42% in PERU MIGRANT, 56.4% in VIANEV). Hypertension, although less prevalent, affected approximately one-fifth of both populations.

### Sex Differences in the Prevalence of Metabolic Disorders

Table 2 provides a detailed view of the distribution of metabolic alterations by sex. Generally, a trend is observed where women have a higher proportion of multiple metabolic alterations than men, especially in the categories of four or more alterations. In the PERU MIGRANT study, this trend is more pronounced, with women consistently representing a higher percentage in the categories of five, six, and seven alterations across all scenarios. In VIANEV, the trend is similar but even more marked. Women represent significantly more in six and seven alterations across all scenarios.

**Table 1.**
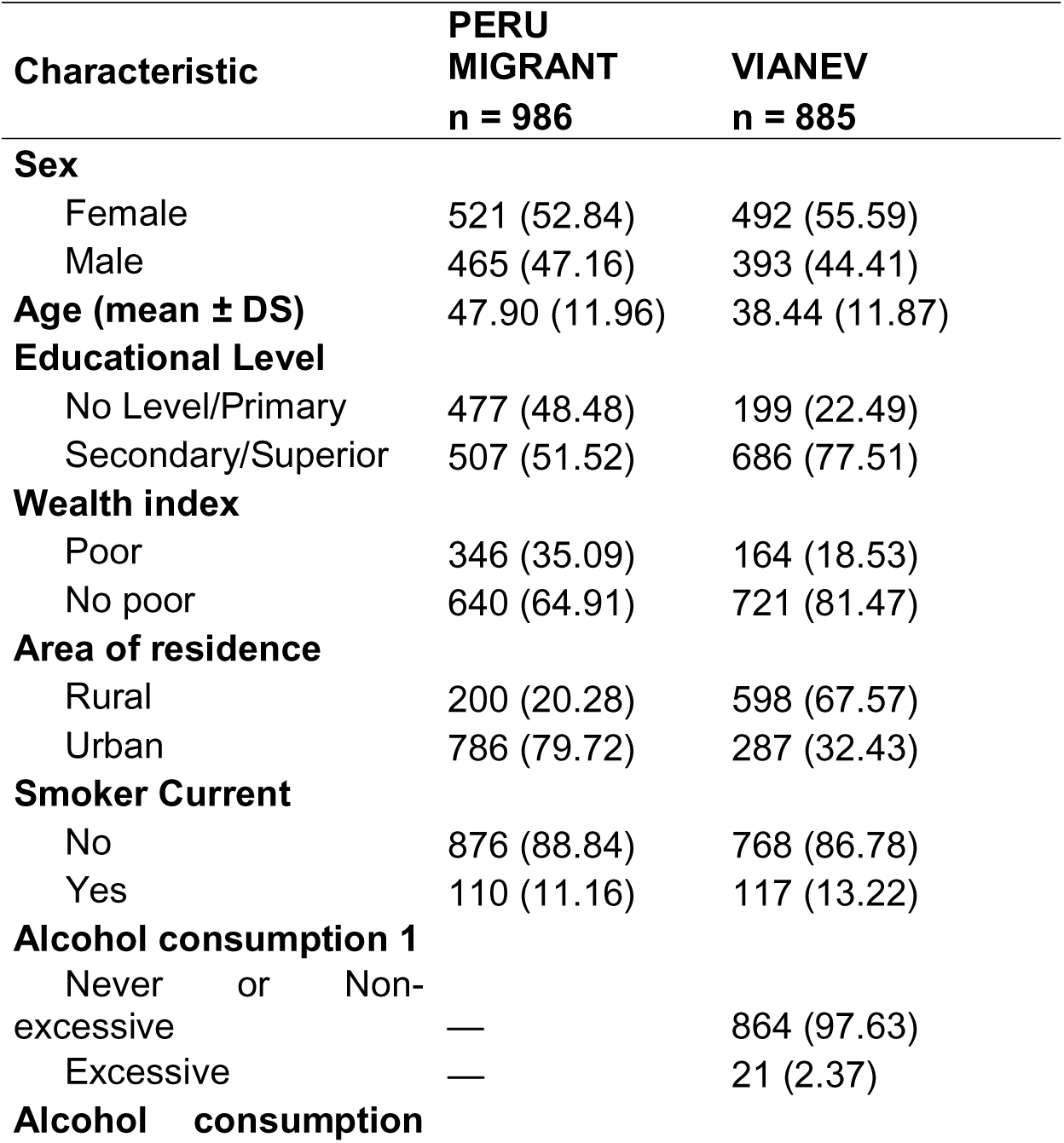
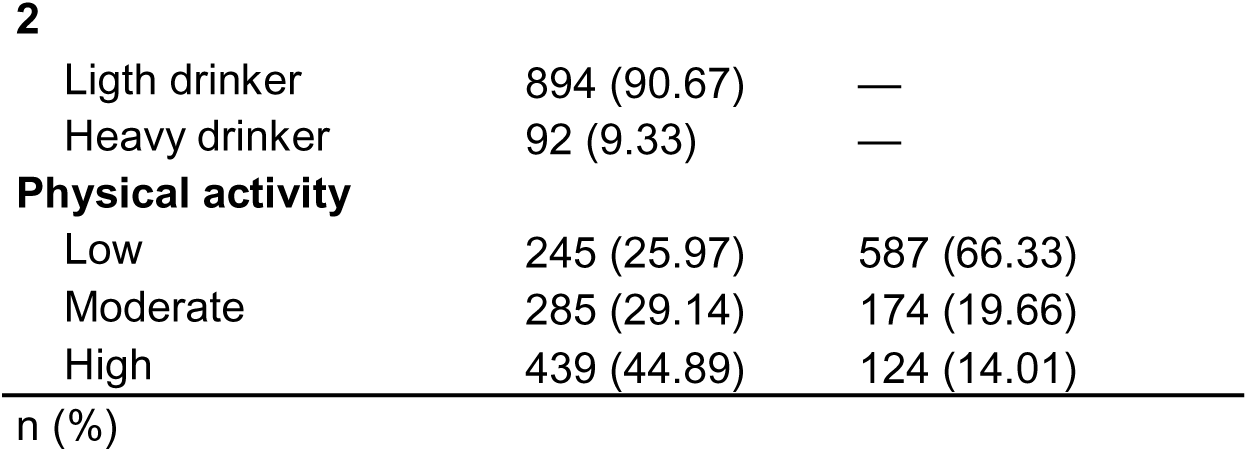
Demographic characteristics of the PERU MIGRANT/VIANEV studies.

**Table 2.** Distribution of metabolic alterations by sex in the PERU MIGRANT/VIANEV studies.

| Metabolic alterations | PERU MIGRANT |  | VIANEV |  |
| --- | --- | --- | --- | --- |
|  | Sex |  |  |  |
|  | Female<br>n (%) | Male<br>n (%) | Female<br>n (%) | Male<br>n (%) |
| **OA-ATP +**<br>**Hyperglycemia-ADA** |  |  |  |  |
| None | 12 (52.17) | 11 (47.83) | 38 (30.16) | 88 (69.84) |
| One | 42 (37.50) | 70 (62.50) | 102 (48.34) | 109 (51.66) |
| Two | 80 (49.69) | 81 (50.31) | 101 (51.53) | 95 (48.47) |
| Three | 74 (48.68) | 78 (51.32) | 98 (52.97) | 87 (47.03) |
| Four | 124 (65.26) | 66 (34.74) | 85 (62.96) | 50 (37.04) |
| Five | 87 (66.92) | 43 (33.08) | 57 (69.51) | 25 (30.49) |
| Six | 54 (71.05) | 22 (28.95) | 32 (86.49) | 5 (13.51) |
| Seven | 11 (57.89) | 8 (42.11) | 6 (75.00) | 2 (25.00) |
| **OA-IDF +**<br>**Hyperglycemia -ADA** |  |  |  |  |
| None | 9 (45.00) | 11 (55.00) | 35 (28.46) | 88 (71.54) |
| One | 33 (32.35) | 69 (67.65) | 80 (44.20) | 101 (55.80) |
| Two | 46 (40.71) | 67 (59.29) | 79 (47.59) | 87 (52.41) |
| Three | 88 (56.41) | 68 (43.59) | 108 (58.38) | 77 (41.62) |
| Four | 138 (65.71) | 72 (34.29) | 101 (64.74) | 55 (35.26) |
| Five | 101 (69.66) | 44 (30.34) | 73 (66.36) | 37 (33.64) |
| Six | 56 (62.22) | 34 (37.78) | 37 (72.55) | 14 (27.45) |
| Seven | 13 (48.15) | 14 (51.85) | 6 (75.00) | 2 (25.00) |
| **OA-ATP +**<br>**Hyperglycemia -OMS** |  |  |  |  |
| None | 17 (40.48) | 25 (59.52) | 38 (29.92) | 89 (70.08) |
| One | 57 (43.18) | 75 (56.82) | 103 (48.36) | 110 (51.64) |
| Two | 85 (50.00) | 85 (50.00) | 104 (52.00) | 96 (48.00) |
| Three | 91 (56.52) | 70 (43.48) | 110 (55.84) | 87 (44.16) |
| Four | 104 (65.00) | 56 (35.00) | 78 (59.54) | 53 (40.46) |
| Five | 84 (66.67) | 42 (33.33) | 55 (72.37) | 21 (27.63) |
| Six | 38 (64.41) | 21 (35.59) | 28 (90.32) | 3 (9.68) |
| Seven | 8 (61.54) | 5 (38.46) | 3 (60.00) | 2 (40.00) |
| **OA-IDF +**<br>**Hyperglycemia -OMS** |  |  |  |  |
| None | 12 (32.43) | 25 (67.57) | 35 (28.23) | 89 (71.77) |
| One | 40 (36.36) | 70 (63.64) | 81 (44.26) | 102 (55.74) |
| Two | 60 (45.45) | 72 (54.55) | 81 (48.21) | 87 (51.79) |
| Three | 113 (62.09) | 69 (37.91) | 120 (61.22) | 76 (38.78) |
| Four | 116 (67.44) | 56 (32.56) | 96 (61.15) | 61 (38.85) |
| Five | 92 (68.15) | 43 (31.85) | 71 (67.62) | 34 (32.38) |
| Six | 43 (55.84) | 34 (44.16) | 32 (76.19) | 10 (23.81) |
| Seven | 8 (44.16) | 10 (55.84) | 3 (60.00) | 2 (40.00) |

Notably, men tend to have a more excellent representation in the categories of none or one alteration, especially in the VIANEV study. Moreover, the distribution appears to be more balanced in both studies’ intermediate categories (two and three alterations). Finally, the differences in distribution between the different criteria (ATP vs. IDF for AO and ADA vs. WHO for hyperglycemia) are evident, but the general trend of higher prevalence of multiple alterations in women is maintained.

Table 3 shows the ordinal logistic regression of the outcome according to sex. In both studies, men are less likely to have metabolic alterations than women (all values are less than 1, with women as the reference), regardless of the criteria used to define metabolic state. Furthermore, the VIANEV study shows a less marked difference between men and women than the other study.

**Table 3.** Ordinal logistic regression for metabolic state disparities according to sex.

| Sex | Metabolic Status |  |  |  |  |  |  |  |
| --- | --- | --- | --- | --- | --- | --- | --- | --- |
|  | OA-ATP +<br>Hyperglycemia-ADA |  | OA-IDF +<br>Hyperglycemia-ADA |  | OA-ATP +<br>Hyperglycemia-OMS |  | OA-IDF +<br>Hyperglycemia-OMS |  |
|  | OR | 95% IC | OR | 95% IC | OR | 95% IC | OR | 95% IC |
| **PERU**<br>**MIGRANT** |  |  |  |  |  |  |  |  |
| Female | Ref. |  | Ref. |  | Ref. |  | Ref. |  |
| Male | 0.44 | 0.35 – 0.55 | 0.40 | 0.32 – 0.51 | 0.45 | 0.35 – 0.56 | 0.41 | 0.32 – 0.51 |
| **VIANEV** |  |  |  |  |  |  |  |  |
| Female | Ref. |  | Ref. |  | Ref. |  | Ref. |  |
| Male | 0.47 | 0.37 – 0.61 | 0.48 | 0.37 – 0.61 | 0.53 | 0.42 – 0.68 | 0.51 | 0.40 – 0.65 |

OR: Odds ratio. CI 95%: Confidence interval at 95%

## Discussion

### Prevalence of Metabolic Alterations

When analyzing metabolic alterations in the population, it is crucial to highlight that, regardless of the criteria used, a small proportion of individuals do not present any alterations. Despite some differences in absolute prevalences, the general distribution pattern of metabolic disorders remained similar across studies. Moreover, while classifications such as altered metabolic state (two or more alterations) or metabolic syndrome (three or more alterations) have been established, it is essential to recognize that each of these metabolic markers carries an independent risk of cardiovascular morbidity and mortality ^(2)^.

This reality raises two fundamental questions: Are we genuinely facing a metabolic epidemic? Is there a problem with the cut-off points used to define these alterations? Particularly concerning is that, even in the most optimistic scenario, using less stringent cut-off points, the prevalence of metabolic alterations (at least one alteration) reaches 87.04% of the studied population. In the worst-case scenario, this figure rises to 87.55%.

These data suggest that, regardless of how metabolic alterations are defined, most populations present at least one metabolism-related cardiovascular risk factor. This situation poses significant challenges for public health and clinical practice, implying that almost 9 out of 10 individuals might require medical intervention or follow-up to prevent future complications ^(3,15,16)^.

These findings could have significant implications for health policies, prevention strategies, and the allocation of healthcare resources. Furthermore, they underscore the need to investigate further the factors contributing to this high prevalence of metabolic alterations in the population. Recent studies have shown that factors such as diet, sedentary lifestyle, and chronic stress play a crucial role in developing these alterations ^(17)^.

This study’s high prevalence of metabolic alterations aligns with global trends. For example, a recent meta-analysis estimated that the worldwide prevalence of metabolic syndrome is approximately 25%, with significant variations between regions and age groups ^(18)^. However, our findings suggest that, when considering individual alterations, the proportion of the affected population could be much higher.

### Healthy Behaviors in Peru

It is essential to consider that healthy behaviors in Peru are significantly diminished, as various studies have demonstrated. Harmful habits such as smoking and alcohol consumption remain at concerning levels or experience marginal reductions. A recent study revealed that the prevalence of smoking in Peruvian adults is 12.2%, while risky alcohol consumption reaches 21.1% ^(19)^.

Particularly alarming is the low consumption of fruits and vegetables in the country. Research has shown that less than 10% of the Peruvian population meets the World Health Organization’s recommendation to consume at least 400 grams of fruits and vegetables daily^(20)^. This deficit in the intake of foods rich in essential nutrients and fiber can significantly contribute to metabolic alterations.

While there is debate about the exact prevalence of metabolic alterations in Peru, the existence of an environmental component that justifies the high observed rates cannot be denied. Factors such as rapid urbanization, changes in dietary patterns towards ultra-processed foods, and increased sedentary behavior have been identified as key contributors to the epidemic of metabolic alterations in developing countries like Peru ^(21)^. Furthermore, recent studies have highlighted the influence of social determinants of health on the prevalence of metabolic alterations in Peru. For example, a significant association has been found between socioeconomic status and the prevalence of metabolic syndrome, being higher in lower socioeconomic groups ^(22)^.

In this context, it is crucial to implement comprehensive public health strategies that address individual risk factors and social and environmental determinants of health. These strategies should include policies to promote healthy eating, increase physical activity, and reduce tobacco and alcohol consumption, adapted to the Peruvian cultural and socioeconomic context ^(23)^.

### Discrepancies in Obesity Criteria

The ATP III criteria recommend higher cut-off points for AO, resulting in a lower prevalence. In contrast, the IDF criteria, which suggest more specific cut-off points for the Latin population, elevate the prevalence to almost 65% of the Peruvian population ^(22)^. This contrast raises crucial questions: Do 65% of Peruvians have obesity? Or should we seek more specific cut-off points for our population?

When examining nutritional status according to BMI, we find that approximately 60% of the Peruvian population is overweight or obese. However, it is essential to note that the prevalence of obesity by BMI is around 26% ^(24)^. This distinction is crucial, as the psychological impact and clinical implications of labeling a patient as “overweight” differ significantly from diagnosing them with “obesity.”

Recent research suggests the need to reevaluate cut-off points for AO in specific populations. A study by Vasquez-Romero proposes that the optimal cut-off points for WC in the Peruvian population could be different from those currently used, which could significantly impact the reported prevalence of AO and metabolic syndrome ^(24)^. Thus, the choice of diagnostic criteria affects epidemiological statistics and has important implications for clinical practice and public health policies.

It is essential to highlight that, regardless of its combination with hyperglycemia, AO, according to IDF criteria, appears to be the main factor responsible for the high prevalence of metabolic alterations. This underscores the need to critically evaluate diagnostic criteria and their applicability in different populations ^(5)^.

Therefore, it is fundamental to reconsider and possibly reclassify the cut-off points used to define AO in Peru, seeking more local and specific criteria. This would improve the accuracy of our epidemiological assessments and allow for more targeted and effective public health interventions.

### Discrepancies in Hyperglycemia

There are also significant discrepancies in diagnostic criteria for hyperglycemia. For years, the ADA has recommended considering values above 100 mg/dl as the cut-off point to define alterations in glucose metabolism. In comparison, the WHO maintains its position of considering values above 110 mg/dl ^(11,12)^.

This divergence creates essential challenges in diagnosing and managing hyperglycemia in patients. Recent studies have found that cardiovascular risk increases significantly only from values above 110 mg/dl, which supports the WHO’s position ^(25)^. This finding has important implications for both clinical practice and public health.

When comparing the prevalences of hyperglycemia according to WHO and ADA criteria, a considerable reduction is observed when using the higher WHO cut-off point. This can be explained by the concentration of a significant number of patients between 100 and 110 mg/dl. This difference is not trivial, as labeling a patient with hyperglycemia not only increases their concerns but can also lead to the application of medical treatments based on medications, which are not exempt from adverse effects.

A systematic review by Richter B et al. found that individuals with fasting glucose between 100-109 mg/dl had a lower risk of progression to type 2 diabetes mellitus than those with levels between 110-125 mg/dl ^(26)^. Furthermore, a meta-analysis by Huang et al. ^(27)^ suggests that optimal cut-off points for prediabetes diagnosis may vary according to ethnicity and geographic region. This highlights the importance of considering population-specific factors when establishing diagnostic criteria.

Thus, it is crucial to consider these implications when diagnosing and treating hyperglycemia. While monitoring glucose levels in all patients is essential, the therapeutic approach may vary. For patients with values between 100-110 mg/dl, interventions could focus more on lifestyle changes, while for those above 110 mg/dl, more aggressive pharmacological interventions could be considered.

Therefore, it is essential to re-evaluate the diagnostic criteria for hyperglycemia, considering not only numerical cut-off points but also the clinical impact and implications for treatment. More specific research on Latin American populations is needed to determine the most appropriate cut-off points and effective management strategies.

### Sex Disparities

Sex disparities in metabolic alterations are a topic of growing interest in epidemiological and clinical research. Our study reveals a consistent trend: Women present a higher prevalence of multiple metabolic alterations than men, regardless of the criteria used to define these alterations.

Our bivariate analysis shows this trend: women are overrepresented in four or more metabolic alterations. At the same time, men tend to have more excellent representation in the categories of none or one alteration. These findings are consistent with recent studies that have found that women tend to present more metabolic alterations than men ^(6)^.

Our results consistently show that women are more likely to present metabolic alterations than men, regardless of the criterion. This finding is consistent with previous studies in Latin America. For example, Márquez-Sandoval et al. ^(28)^ found a higher prevalence of metabolic syndrome in women in several Latin American countries.

Moreover, hormonal differences and body composition between men and women could influence metabolic risk. A study by Mauvais-Jarvis and Hevener ^(29)^ highlights how estrogens affect glucose and lipid metabolism differently in women. On the other hand, differences in gender roles, access to healthcare, and physical activity patterns could contribute to these disparities. A study by Creber in Peru found that women had lower physical activity than men, which could increase their metabolic risk ^(30)^.

Our findings and existing evidence underscore the need for a sex-differentiated approach to evaluating and managing metabolic alterations. More research is needed, particularly in Latin American populations, to better understand the underlying reasons for these disparities and to develop prevention and treatment strategies considering these differences between sexes.

### Importance for Public Health

This study is of fundamental importance for public health in Peru and internationally. By revealing the high prevalence of metabolic alterations in the Peruvian population, it provides a solid basis for formulating more effective and targeted public health policies. Identifying discrepancies in diagnostic criteria and their implications for the reported prevalence of abdominal obesity and hyperglycemia underscores the need to re-evaluate and possibly adapt these criteria to the specific characteristics of the Peruvian and Latin American populations in general.

At the international level, this study contributes to the growing body of evidence suggesting the need for population-specific diagnostic criteria. This is particularly relevant in an increasingly globalized world, where health recommendations are often applied universally without considering ethnic and regional variations. The findings of this study can influence how international guidelines for diagnosing and managing metabolic alterations are developed and used, promoting a more personalized and culturally sensitive approach in preventive medicine and global public health.

### Conclusions and Recommendations

This study reveals a high prevalence of metabolic alterations in the Peruvian population, with significant variations according to the diagnostic criteria. The observed discrepancies in the prevalence of AO and hyperglycemia, depending on the cut-off points employed, underscore the need to re-evaluate these criteria for the Peruvian population and possibly for other Latin American populations. Furthermore, the observed differences between sexes in the prevalence of metabolic alterations, particularly in abdominal obesity, suggest the need to consider sex-differentiated approaches in prevention and management strategies.

The findings of this study have important implications for clinical practice and public health policies. Even with more conservative criteria, the high prevalence of metabolic alterations indicates an urgent need for preventive interventions at the population level. However, the variability in diagnoses according to the requirements also suggests the importance of a more nuanced and personalized approach in assessing and managing individual metabolic risks.

Additional studies are recommended to determine the optimal cut-off points for abdominal obesity and hyperglycemia in the Peruvian population, considering ethnic, regional, and sex differences. Meanwhile, at the public health policy level, it is recommended to implement comprehensive strategies that address metabolic risk factors from multiple angles. This includes public education campaigns on healthy lifestyles, policies that promote balanced nutrition and physical activity, and improvements in access to preventive health services. Additionally, it is suggested that national clinical guidelines be developed that consider this study’s findings, guiding health professionals on how to interpret and manage metabolic alterations in the specific context of the Peruvian population.

## Authors’ contribution

**Víctor Juan Vera-Ponce:** Conceptualization, Investigation, Methodology, Resources, Writing - Original Draft, Writing - Review & Editing

**Luisa Erika Milagros Vásquez-Romero:** Investigation, Project administration, Writing - Original Draft, Writing - Review & Editing

**Joan A. Loayza-Castro**: Investigation, Resources, Writing - Original Draft, Writing - Review & Editing

**Fiorella E. Zuzunaga-Montoya:** Software, Data Curation, Formal analysis, Writing - Review & Editing

**Jhonatan Roberto Astucuri Hidalgo:** Validation, Visualization, Writing - Original Draft, Writing - Review & Editing

**Carmen Inés Gutierrez De Carrillo:** Methodology, Supervision, Funding acquisition, Writing - Review & Editing

## Acknowledgments

A special thanks to the members of Universidad Nacional Toribio Rodríguez de Mendoza de Amazonas (UNTRM), Amazonas, Peru for their support and contributions throughout the completion of this research.

## Financial Disclosure

This study was financed by Vicerectorado de Investigación de la Universidad Nacional Toribio Rodríguez de Mendoza de Amazonas.

## Conflict of interest

The authors declare no conflict of interest.

## Informed consent

The primary’s study’s from which the database was obtained provided the required informed consent, however, for the present study it was not required.

## Data availability

The data supporting the findings of this study can be accessed by the original research paper at the following link:

- PERU MIGRANT: https://figshare.com/articles/dataset/PERU_MIGRANT_Study_Baseline_dataset/3125005
- VIANEV: https://www.datosabiertos.gob.pe/dataset/estado-nutricional-en-adultos-de-18-59-a%C3%B1os-per%C3%BA-2017-%E2%80%93-2018

## Notes

### Competing Interest Statement

The authors have declared no competing interest.

### Summary of Updates

In this new version, the order of the final sections of the article was modified, adding after the conclusions, the contribution of authors, the acknowledgments and the financing, which were also modified, continuing with the conflicts of interest, the informed consent, which was also modified, the capacity of the data and finally the bibliography.

